# Critical success factors for high routine immunization performance: A case study of Senegal

**DOI:** 10.1101/2022.01.25.22269847

**Authors:** Zoe Sakas, Kyra A. Hester, Katie Rodriguez, Saly Amos Diatta, Anna S. Ellis, Daouda Malick Gueye, Dawn Matapano, Pr Souleymane Mboup, Emily Awino Ogutu, Chenmua Yang, Robert A. Bednarczyk, Matthew C. Freeman, Moussa Sarr, the Vaccine Exemplars Research Consortium

**Affiliations:** Rollins School of Public Health, Emory University, Atlanta, GA, USA; Need to ask for Katie’s affiliation; Institut de Recherche en Santé de Surveillance Epidemiologique et de Formation, Dakar, Senegal; College of Arts and Sciences, Emory University, Atlanta, GA, USA; H. Milton Stewart School of Industrial and Systems Engineering, Georgia Institute of Technology, Atlanta, GA, USA; Need to ask for Bonheur’s affiliation; Emory University School of Medicine, Emory University, Atlanta, GA, USA; Biden School of Public Policy and Administration, University of Delaware, Newark, DE, USA; College of Engineering, Georgia Institute of Technology, Atlanta, GA, USA; School of Public Policy, Georgia Institute of Technology, Atlanta, GA, USA

## Abstract

**BACKGROUND:** The essential components of a vaccine delivery system are well-documented, but robust evidence is lacking on how policies and implementation strategies are operationalized to drive catalytic improvements in coverage. To address this gap, we identified success factors that supported improvements in routine immunization coverage in Senegal, especially from 2000 to 2019.

**METHODS:** We identified Senegal as an exemplar in the delivery of childhood vaccines through analysis of DTP1 and DTP3 coverage data. Through interviews and focus group discussions at the national, regional, district, health facility, and community-level, we investigated factors that contributed to high and sustained vaccination coverage. We conducted a thematic analysis through application of implementation science frameworks to determine critical success factors. We triangulated these findings with quantitative analyses using publicly available data.

**RESULTS:** The following success factors emerged: 1) Strong political will and prioritization of resources for immunization programming supported urgent allocation of funding and supplies; 2) Collaboration between the Ministry of Health and Social Action and external partners fostered innovation, capacity building, and efficiency; 3) Improved surveillance, monitoring, and evaluation allowed for timely and evidence-based decision making; 4) Community ownership of vaccine service delivery supported tailored programming and quick response to local needs; and 5) Community health workers spearheaded vaccine promotion and demand generation for vaccines.

**CONCLUSION:** The vaccination program in Senegal was supported by evidence-based decision making at the national-level, alignment of priorities between governmental entities and external partners, and strong community engagement initiatives that fostered local ownership of vaccine delivery and uptake. High routine immunization coverage was likely driven by prioritization of immunization programming, improved surveillance systems, a mature and reliable community health worker program, and tailored strategies for addressing geographical, social, and cultural barriers.

## 1. Introduction

Vaccination is recognized as one of the most influential public health interventions of the last century, averting an estimated 2-3 million deaths annually [1-3]. The Global Vaccine Action Plan (GVAP) targeted least 90% country-level coverage of the third dose of diphtheria, tetanus, pertussis vaccine (DTP3) among 1-year-old children, a globally recognized proxy for vaccination system performance, and 80% DTP3 coverage for subnational areas [4]. By 2018, only 95 of the 193 World Health Organization (WHO) Member States achieved the GVAP targets [5]. The African Region reported the lowest DTP3 coverage at 74% in 2019, a slight improvement from 71% coverage in 2010 [6]; however, Senegal drastically improved its DTP3 coverage from 52% to 93% between 2000 and 2019 [6, 7]. Examining the success of the vaccination program in Senegal provides an opportunity to identify and describe critical factors for effective vaccine systems.

The essential components of an effective vaccine delivery system are well-established and include strong governance and leadership, healthcare financing, human resources, a robust supply chain, and information systems [8]. Previously identified determinants of vaccination coverage include intent to vaccinate, community access, and health facility readiness [9]. However, evidence is lacking on how policies and implementation strategies are operationalized - through strong governance and financing mechanisms - to drive and sustain catalytic changes in coverage.

The purpose of this study was to identify critical success factors that contributed to exemplary growth in childhood routine immunization coverage in Senegal. Findings from this research may identify transferable lessons and support actionable recommendations to improve national immunization coverage in other settings [10]. Many countries have seen stalled progress in the last decade, so identifying the factors and mechanisms for accelerating progress could be critical for expanding vaccination program reach and retention. We applied a positive deviance approach through investigating vaccine delivery systems in exemplar countries [10-12]. Here we describe the design, adaptation, and implementation of successful policies and programs in Senegal.

## 2. Methods

We employed a qualitative case study design to investigate factors that contributed to high and sustained vaccination coverage in Senegal through key informant interviews (KIIs) and focus group discussions (FGDs) at the national, regional, district, health facility, and community-level. We triangulated these findings with quantitative analyses using publicly available data, which are published elsewhere and referenced throughout this paper [11, 12]. This study was nested within the Exemplars in Vaccine Delivery project [10].

Prior to data collection, we developed a conceptual model (Figure 1), to organize factors that impact childhood vaccination coverage globally. This model was based on the work of Phillips et al. and LaFond et al. alongside a broader review of the vaccine confidence and coverage literature [9, 13].

**Figure 1.**
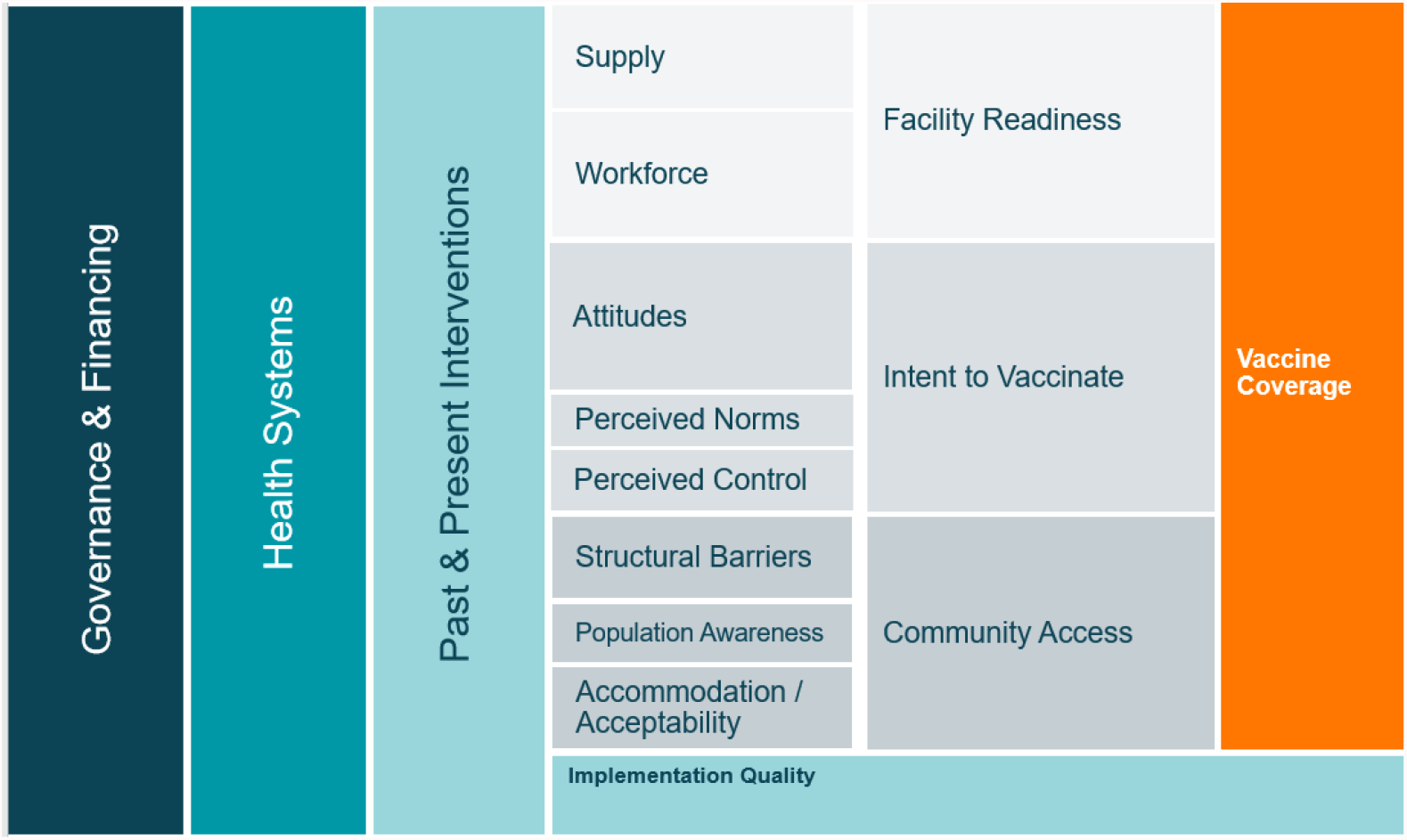
Conceptual model of the drivers of vaccination coverage.

### 2.1. Study setting

Senegal was selected as an exemplar in vaccine delivery based on DTP1 and DTP3 coverage estimates which served as proxies of the vaccine delivery system, with DTP1 as an indicator for access and DTP3 as an indicator for continued use of immunization services [11, 14]. Senegal is in Sub-Saharan Africa and is currently classified as a lower-middle-income country with a growing economy based on agriculture, fishing, and tourism [15]. The MoH in Senegal consists of three levels: the national-level, medical regions, and health districts. There are 14 medical regions and 77 health districts (as of 2019), covering more than 1,859 health posts [16].

In consultation with national stakeholders and available data, we selected three regions in Senegal as study locations while considering heterogeneity via the following characteristics: 1) contextual factors including population density, ethnicity, and geography; 2) high and low vaccine performance, grouped by >80% and ≤80% DTP3 coverage; and 3) inclusion of the capital city of Dakar. Ziguinchor, Dakar, and Tambacounda regions were ultimately selected with DTP3 coverage of 93.6%, 92.6%, and 77.0% in 2017, respectively. We selected three districts within each region, including: Ziguinchor, Oussouye, and Diouloulou in Ziguinchor; Rufisque, Mbao, Keur Massar in Dakar; and Tambacounda, Koupetoum, and Goudiry in Tambacounda.

### 2.2. Quantitative data collection and analysis

Quantitative data were collected through secondary datasets, which included information from the Ministry of Health and Social Action (referred to as MoH throughout) and other partners. Data were used to estimate routine immunization coverage from 2000 to 2019 and to uncover trends related to improvements and sustainability. Additional analyses were conducted to identify indicators that may be associated with immunization coverage success among low- and lower-middle-income countries using cross-country and multi-year mixed-effects regression models to statistically test financial, development, demographic, and other country-level indicators. The results from these analyses will be presented in a forthcoming paper. These quantitative analyses informed the qualitative findings presented here through providing additional context.

### 2.3. Qualitative data collection and analysis

Qualitative data were collected between December 2020 and April 2021 at the national, regional, district, health facility, and community levels. The interview guides were informed by the Consolidated Framework for Implementation Research (CFIR) [17] and the Context and Implementation of Complex Interventions (CICI) framework [18]. Key informant interview (KII) and focus group discussion (FGD) guides were translated in French and Wolof languages by research assistants. All interview guides were piloted before use and adjusted iteratively throughout data collection. An initial list of KIIs was developed with local research partners and MoH officials; snowball sampling was used to identify additional key informants. Our sampling approach aimed to include a diverse sample of participants in regard to geographic location and demographic qualities. Caregivers and community health workers (CHWs) were recruited for FGDs from health facility catchment areas with the assistance of local health staff. The duration of KIIs and FGDs averaged one and a half hours. KIIs and FGDs were audio-recorded with the permission of participants. Research files, recordings, and transcriptions were de-identified and password protected. The activities are summarized in Table 1.

**Table 1.** Research activities completed from December 2020 to April 2021.

| Method | Participant | Number of activities |
| --- | --- | --- |
| **Key informant interviews (KIIs)** | Ministry of Health at the national level | 4 |
|  | Ministry of Education at the national level | 1 |
|  | Association of Journalists for Health (AJSP) | 1 |
|  | Partner organizations | 3 |
|  | Regional health office | 7 |
|  | District health office | 38 |
|  | Head nurses in charge of health facilities | 6 |
|  | Community leaders | 2 |
| **Focus group discussions (FGDs)** | Community workers and volunteers | 10 |
|  | Mothers | 9 |
| **Total** | n/a | **81** |

A theory-informed thematic analysis of the transcripts identified success factors for the immunization program. We developed a codebook using a deductive approach, applying constructs from the CFIR and CICI frameworks and adjusted the codebook based on emerging themes. Transcripts were coded and analyzed using MaxQDA2020 software (Berlin, Germany). All transcripts were coded, relevant themes were identified, and visual tools were used to illustrate the findings. We considered setting and participant roles while identifying key points, and further contextualized data using historical documents and a literature review. Topic guides and codebooks used for data collection and analysis can be found on our Open Science Framework webpage [19].

### 2.4. Ethical approval

The study was reviewed and approved by the National Ethical Committee for Health Research (CERNS; Comité National d’Ethique pour la Recherche en Santé) in Dakar, Senegal (00000174). This study was also approved as exempt human research by the Institutional Review Board committee of Emory University, Atlanta, Georgia, USA (IRB00111474). All participants provided written consent. One 12-year-old girl was recruited for and participated in a FGD with mothers, and although she was accompanied by a guardian, we did not use data from her for the analysis due to her young age. Two 15-year-old girls were also recruited for FGDs with mothers, which we did use for analysis; they were both accompanied by their guardians who also provided verbal consent for them to participate in the study.

## 3. Results & Discussion

KIIs were conducted at the national (n=9), subnational (n=52), and community (n=2) levels. FGDs were conducted with community health workers (n=10) and mothers (n=9) within the nine selected districts. Demographic information for 109 of the FGD participants can be found in Table 2.

**Table 2.** Demographic information for FGD participants in Senegal.

| Characteristics, mean (range) | CHWs | Mothers | Total |
| --- | --- | --- | --- |
| Number of participants | 47 | 62 | 109 |
| Age of participants | 41 (22-70) | 27 (15-40) | 33 (12-70) |
| Number of children | 4 | 3 | 3 |
| Age of youngest child vaccine decisions are made for | 5 | 2 | 3 |
| **Highest level of education, n (%)** |  |  |  |
| No formal schooling | 5 (11) | 7 (11) | 12 (11) |
| Some schooling, less than 5 years | 11(24) | 11 (18) | 22 (20) |
| Some schooling, b/w 6-11 years | 15 (33) | 36(58) | 51 (47) |
| High school completed | 15(33) | 5 (8) | 20 (19) |
| Attended University | 0 (0) | 2 (3) | 2 (2) |
| Other | 0 (0) | 1 (2) | 1 (1) |
| **Religion, n (%)** |  |  |  |
| Islam | 40(85) | 51 (82) | 91 (83) |
| Catholicism | 7 (15) | 10 (16) | 17 (16) |
| Protestantism | 0 (0) | 1 (2) | 1 (1) |

### 3.1. Routine immunization performance in Senegal

To assess the performance of the vaccine delivery system in Senegal, we used coverage estimates for BCG, DTP1, DTP3, MCV1, and Pol3 from WUENIC [7, 20] and estimates for full coverage of all 8 vaccine doses (3 doses each of DTP and Pol, 1 dose each of BCG and MCV) from UNICEF [20]. According to this analysis, coverage for these routine immunizations in Senegal steadily increased from 2002 to 2007 and remained relatively constant from 2007 to 2019.

Through our literature and document review, we identified policies, programs, and events that may have impacted vaccination coverage in Senegal from 2000 to 2019, which are illustrated in Figure 4 alongside coverage estimates. Notably, in 2001 Senegal instituted an 8-year strategic plan for eliminating measles [21], a new constitution guaranteeing the right to health [22], and a national expansion of their Integrated Management of Childhood Illness (IMCI) program (with a community-based expansion in 2006). Additionally, Gavi support for Senegal began in 2002, which correlates with the sharp increases in coverage from 2002 to 2007.

There was a decrease in reported coverage for all vaccines in 2018, coinciding with healthcare worker strikes that took place from roughly April 2018 to January 2019. Senegal has experienced numerous healthcare worker strikes in the past few decades. During the strikes (particularly in 2018), healthcare workers from all levels refused to collect data on their patients, withheld data from the MoH and partners, and sometimes even restricted vaccination services [23-25].

### 3.2. Development of a conceptual framework to highlight mechanisms for successful vaccine delivery in Senegal

Success factors for the Senegalese vaccine delivery system represent both homegrown innovations and adaptations of global guidelines to the local context. Through qualitative analysis, we focused on how policies were formalized and operationalized to support high and sustained routine immunization coverage in Senegal, as well as how programming was shaped by local context and culture.

Our analysis led to the development of an explanatory framework that aims to categorize mechanisms of success that likely contributed to high and sustained vaccination coverage in Senegal. Although existing literature describes the requirements for successful vaccine delivery (Figure 2) [9], there is a lack of evidence on how governance structures and health systems function within successful programs. Figure 5 illustrates this framework and highlights the key drivers as they relate to Senegal’s immunization system. We found these mechanisms contributed to successful vaccine delivery between and within the levels of implementation in Senegal. The functional definitions for these mechanisms and levels of implementation are found in Table 3.

**Table 3.**
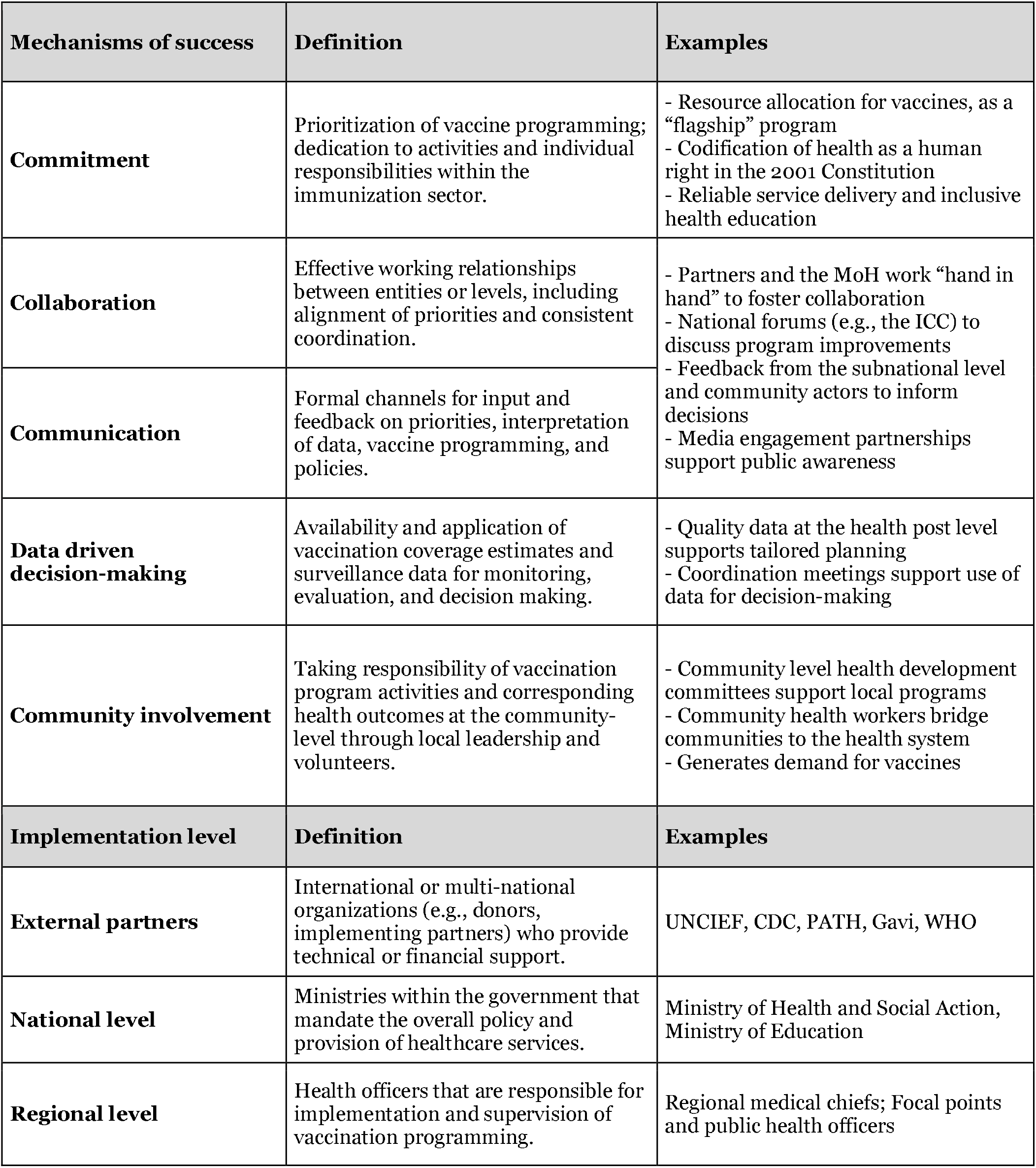
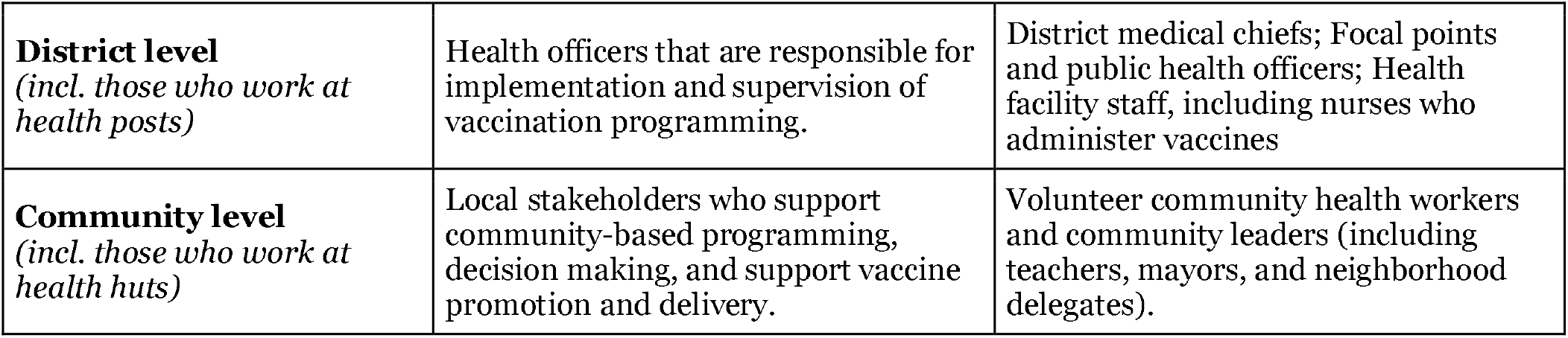
Functional definitions for featured mechanisms of successful vaccine delivery and their implementation levels.

**Figure 2.**
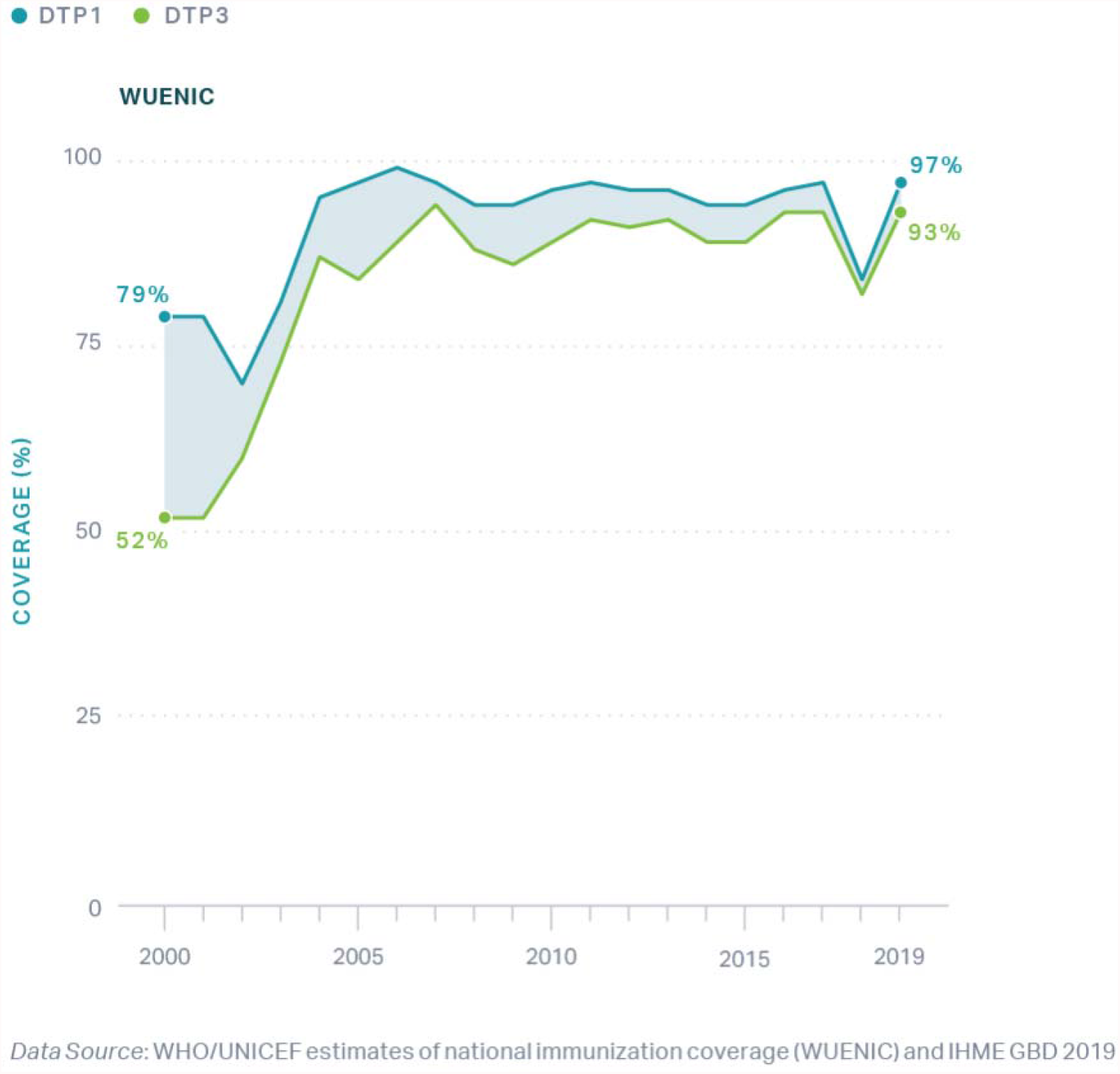
National-level WHO UNICEF Estimates of National Immunization Coverage (WUENIC) for DTP1 and DTP3 in Senegal, 2000 – 2019.

**Figure 3.**
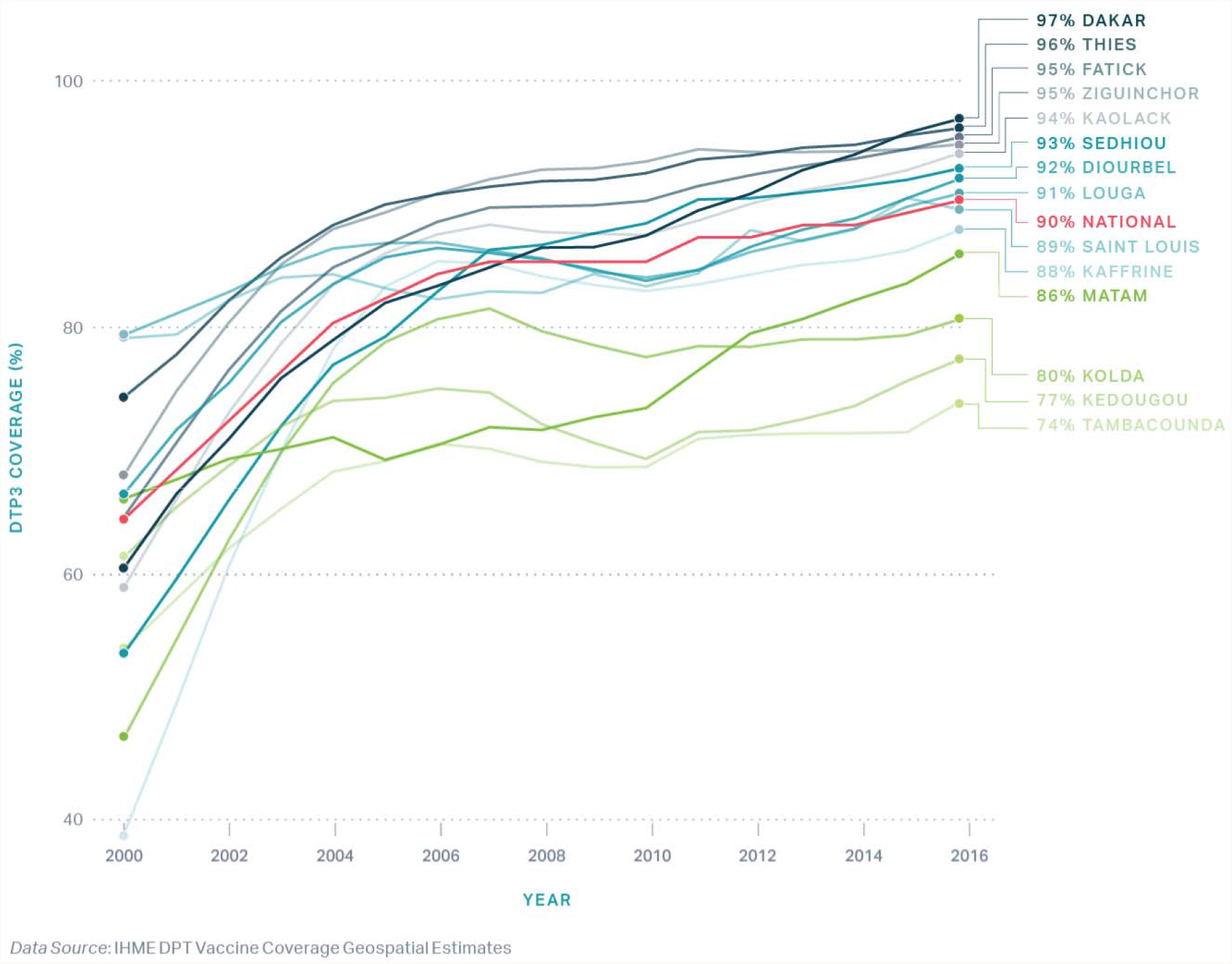
DHS DTP3 coverage in Nepal, by Province, 2000 – 2016.

**Figure 4.**
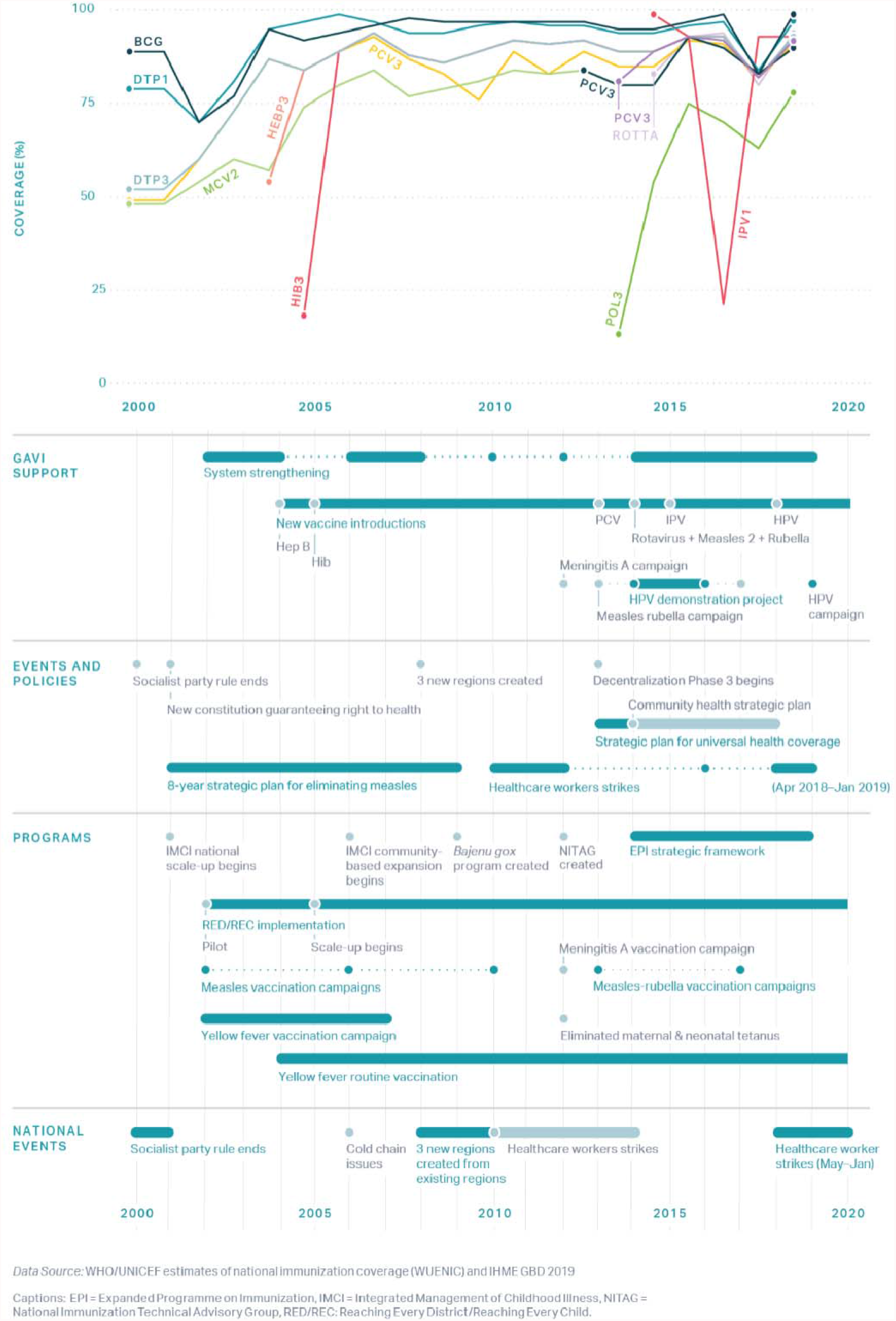
Routine immunization coverage in Senegal annotated with key events, programs, and policies from 2000 to 2019.

**Figure 5.**
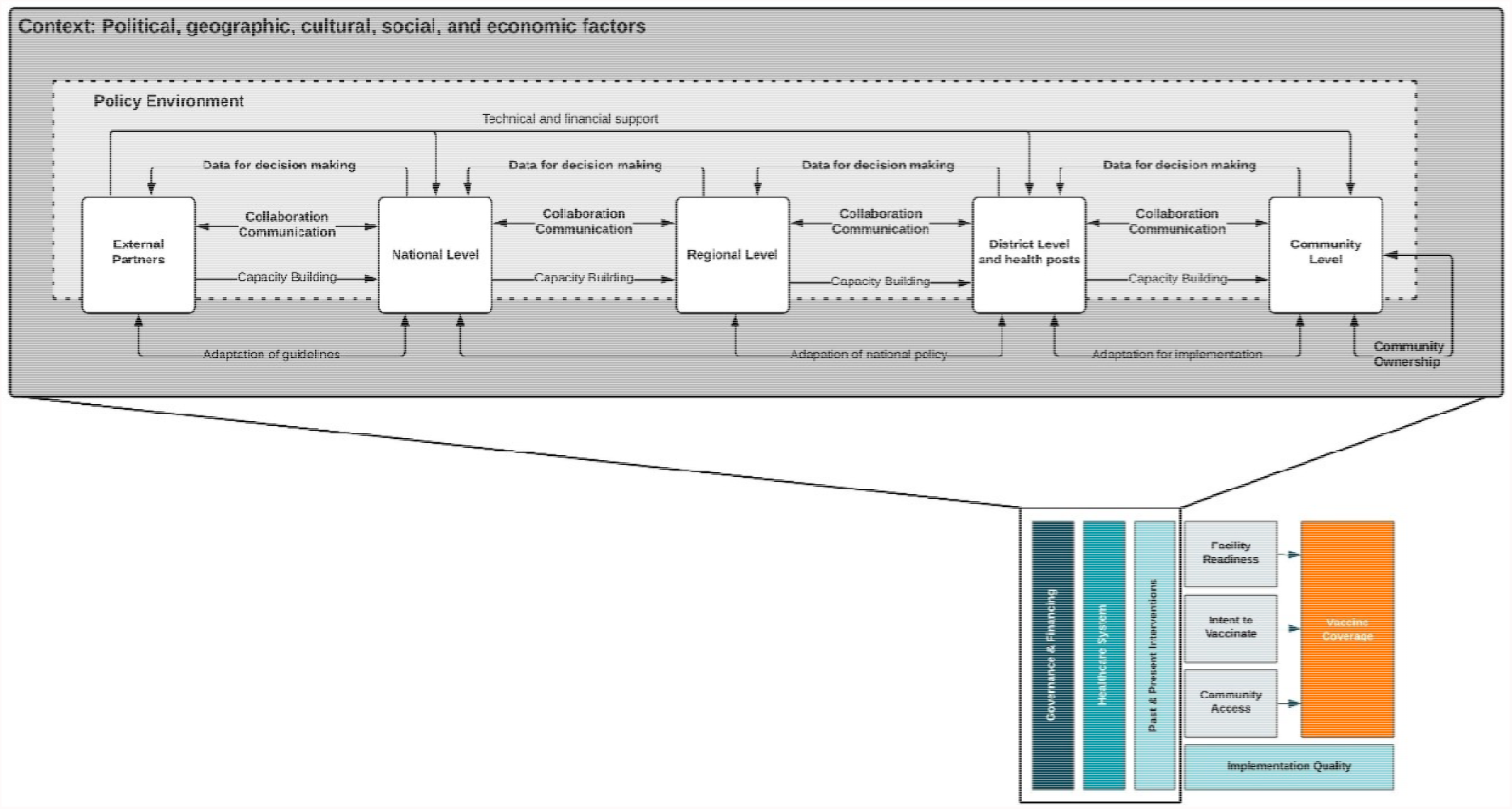
Critical factors and governance mechanisms that supported high coverage of routine vaccinations in Senegal.

This framework illustrates critical mechanisms that contributed to successful vaccine delivery in Senegal according to key informants. The purpose of this framework is to provide a road map for the key findings from this study, which are described in the sections below. As depicted, the policy making environment – which is composed of the MoH at multiple levels and external partners – sits within the context of Senegal, including political, economic, cultural, and geographic factors. Mechanisms for governance and decision-making and how they operate between and within levels of the health system are represented by the arrows. Commitment to achieving equitable health outcomes supports the execution of all mechanisms within the framework. The functional definitions for these mechanisms and levels of implementation are found in Table 3. Similar frameworks have been developed for other exemplar country case studies, including Nepal and Zambia [26, 27].

### 3.3. Commitment mechanism

#### 3.3.1. Prioritization of vaccine programming to reduce childhood mortality

Childhood vaccination is considered a priority, or “flagship program” by key informants because: 1) it is preventative, rather than curative; 2) there has been a tangible and visible decrease in childhood mortality; 3) it is the “right” of all children in Senegal; and 4) “children are the future” of the country and should be protected. MoH staff, external partners, service providers, and community members all described their commitment to the vaccine delivery system and cited that the prioritization of vaccine programming greatly supported catalytic improvements in coverage. In Senegal, vaccine programming is prioritized for policy development, external partner engagement, strategic planning of health-related programs and activities, and both internal and external funding.

> *“What motivated me to do this activity was that it is an activity of prevention… And since the children are a vulnerable group, it is better to focus on prevention than to wait until the children are sick…When we focus on immunization, we improve in the fight for good health of the population.” (Head Nurse, Ziguinchor)*

Service providers and mothers acknowledged vaccines as a right for all children and a priority within their communities. Service providers - including volunteer community health workers, nurses, and other health personnel – noted the importance of reliable, consistent, and equitable service delivery, alongside the promotion of vaccines and inclusive health education, in order to protect children in their communities. Similarly, mothers noted that they prioritize vaccination over other household and business activities to ensure their children are protected from disease.

> *“Vaccination is to fight against diseases, it is prevention, this is the flagship program. It is our duty as service providers, as nurses. The love that we had from the start is what really helps us do the work with respect. We know that [vaccines] give us concrete results.” (Supervisor, Tambacounda)*

> *“When going to vaccinate my child, I cancel all my activities and leave instructions at home to be able to devote myself to the vaccination. Even if I have to go back late, I prefer to stay at the [health facility] until he receives his vaccine.” (Mother, Dakar)*

#### 3.2.2. Health as a human right is codified in the Constitution

Health (including vaccines) is considered a human right in Senegal, which implies that the government is the responsible duty-bearer for service provision and allocates responsibility to parents to vaccinate their children. In 2001, the Constitution of Senegal codified health as a right - as stated in article #14: “the state and the public authorities have the social duty to watch over the physical, mental, and health of the family” [22] - which illustrated the commitment of the national-level government to improving the health system.

> *“It is valued first by making vaccination a right for every child, and to explain that without vaccination children are at risk. We made it possible to put mothers and caregivers at the heart of their own health, of their own children. When you don’t explain to the mother that vaccination is very important, it can’t be valued, so there is an involvement, there is adherence, and inclusion of the community in programming.” (Focal point, Ziguinchor)*

A National Health Strategic Plan (NHSP) is developed for every ten-year period in order to sustain decision-making through different administrations. Decision-makers refer to existing strategic plans during coordination meetings between the MoH, external partners, and technical working groups.

### 3.4. Collaboration and communication mechanisms

#### 3.4.1. Partners and the MoH work “hand in hand” to foster open communication

Close collaboration – including open communication and reliable coordination – between the MoH and external partners was essential for decision-making, funding allocation, and implementation of policies and programs at national and subnational levels. Distinct roles of partners at the national level allowed for effective decision-making. Each partner (e.g., WHO, UNICEF, PATH) was recognized for its specific field of intervention and would lead policy development and program implementation for that component of the vaccine system. With their different fields of expertise, partners and the MoH would work together to ensure successful vaccination programming, with the MoH having the final say for decision-making at the national level.

> *“When we have a strategic document to work on, the partners get involved according to their area of intervention*… *Although each one of us has a specific field of intervention, we pull together when it comes to implementing and monitoring the program. That’s how policy documents are designed. In this teamwork, you cannot tell who the partner is or who is from the Ministry of Health, because we work hand in hand. Because we are all in the same boat, we want to have success*…*” (External partner at the national level)*

The policy environment in Senegal is led by national-level stakeholders within the MoH – relied heavily on the application of international guidelines and policies, which are adapted to meet context-specific priorities and challenges. Key informants at the national, regional, and district levels reported referring to international guidelines during coordination meetings, discussing them as a team, and determining how they may be implemented in Senegal. Examples of international guidelines that were implemented in Senegal include health systems strengthening, Reaching Every District/Child (RED/REC), introduction of new vaccines, and cold chain expansion [28-30]. Evaluations conducted by external partners also influenced strategic planning as the MoH may prioritize these goals or targets to maintain external partner support and resources.

> *“Nowadays, you cannot develop a policy without taking the international guidelines into account. We provide these international guidelines in order to see what is being done elsewhere in the world. Based on that, they define their strategies, their plans.” (External partner at the national level)*

At the national level, the Inter-agency Coordinating Committee (ICC) functions as a forum to support evidence-based decision-making, open collaboration between the MoH and external partners, and policy development [31]. During ICC meetings, members discuss how to improve programming and resource allocation to meet challenges determined by monitoring and evaluation of surveillance data. Existing health policies, including the current NHSP, are referred to during meetings to ensure that priorities are continuous and aligned. One key informant spoke to the informal nature of the ICC, explaining that it is “almost a second home” due to the open and fluid communication between members. This relationship is in part due to the regularity of communications between entities – whether in meetings, by phone, or online.

#### 3.4.2. Feedback from stakeholders at all levels enables effective collaboration

Decision-making for resource allocation relies on feedback from subnational and community actors via bottom-up and top-down mechanisms. For example, subnational levels (e.g., regions, districts, and health posts) create tailored strategies and communication plans to reach the local population, which are reviewed and validated at the national level. With the advent of decentralization, subnational levels retain the capacity to tailor local initiatives to their respective environment.

External partners also work directly with subnational levels, including district health offices and health posts. Subnational partner engagement varies by region and district and may circumvent the national level for efficiency, with approval from the MoH. For example, districts may share their strategic plans with external partners directly, who then validate and provide support according to the needs of the district – this is supported by RED/REC policies at the national level. External partners support financing, supervision, monitoring, evaluation, and implementation of strategies, including technical advisory for regional and district-level health officers, development of health education materials, and construction of health huts. Subnational MoH officers mentioned that innovation from external partners pushed them to adapt strategies to the local context and respond to challenges.

#### 3.4.3. Media engagement partnerships support public awareness

Collaboration between the MoH and media organizations supported the dissemination of accurate health information to communities that aligned with the priorities and programs of the MoH. At the national level, the Association of Journalists for Health (*L’Association des Journalistes en Santé, Population et Développement*, referred to as AJSP) is an independent group of media experts – including members from TV, radio, written press, and online media – that focuses on topics within the health and development sectors. According to key informants, the AJSP developed a “strategic partnership” with the MoH in order to effectively create awareness and dispel rumors at the community level [32]. Press caravans (defined as small-scale campaigns often at the community level) involve AJSP members visiting communities to disseminate information on health topics and to learn from community members about challenges on the ground. Journalists who are part of the press caravan are trained by the MoH and mobilized to areas that have experienced success in order to gain information about effective programming; or to areas that have experienced challenges in order to share lessons learned.

At the district level, health officers engage local media outlets to promote vaccines. Collaboration with local radio channels was mentioned by participants in all regions assessed for this study (Dakar, Ziguinchor, and Tambacounda). Stations may provide airtime for vaccination-specific broadcasts or commercials for the MoH. Vaccination is not the only health topic covered by the media – depending on the needs of the district, health officers assign different topics to media personnel, including child health, vaccination, and sexual and reproductive health. Key informants mentioned that peer-to-peer learning, facilitated by the radio, supported demand generation in their communities.

> *“Here, we already have only one radio. It is community radio, which allows us to broadcast. They are really our partners because they have opened a branch every week to talk about health problems. So, this radio program is really on Sunday evening, but it can happen that we use another day. And it really allows us to communicate with the population for free.” (Focal point, Ziguinchor)*

### 3.5. Data driven decision-making mechanism

#### 3.5.1. Quality data at the health post level supports tailored strategies

The Senegalese health system utilizes District Health Information Software (DHIS2) for data management, coordination, supervision, and analysis at the national, regional, and district health offices, and health posts. Electronic data management software in health posts supports improvements in data quality at the local level compared to paper tracking. Head nurses at health posts submit weekly data monitoring reports through the DHIS2 and district focal points aggregate and validate the data. The district health office provides feedback to the health posts through DHIS2 and helps develop tailored strategies as needed to reach missing children and improve coverage.

Key informants discussed the importance of a timely, reliable, and accurate vaccination coverage surveillance system. Currently, there are two surveillance divisions within the MoH - the Directorate of Prevention and the Center for Health Emergency Operations [33]. The Division of Surveillance and Vaccine Response was established within the Directorate of Prevention. Specific roles and departments for surveillance operations allow for specialization and expertise from the national to district level. Surveillance data illustrate coverage gaps and highlight if some areas or populations might require additional investments to support evidence-based decision-making.

> *“The surveillance data allows us to see if all the diseases that are in the EPI are actually eradicated in the area or are in the process of being eradicated. Because if we vaccinate it proves that we should not have this disease. Therefore, the surveillance makes it possible to follow, to see whether, despite the vaccination, there are cases of diseases that are monitored in the EPI.” (Supervisor, Tambacounda)*

#### 3.5.2. Microplanning cultivates tailored programming

Microplanning at the local level, through the RED/REC approach, supported tailored outreach strategies for diverse populations. As identified by regional and district key informants, the strengthening and implementation of microplans through the RED/REC policy guidelines was a contributing factor for successful vaccine delivery and uptake. Bi-monthly district meetings introduced by PATH allowed for greater engagement of district-level personnel and head nurses of local health posts to monitor vaccination coverage and coordinate outreach strategies [34, 35]. Community-level microplanning occurs at monthly health post meetings, which include head nurses, volunteer community health workers, and community leaders.

Outreach strategies are implemented to reach all households, especially those in remote areas or far from health posts or other health facilities. Health huts, which are typically constructed by external partners and maintained by the community members, are used for awareness generation activities, sensitization, and community mobilization. Outreach strategies may occur at health huts or elsewhere, and they can be “advanced” (5-15km from the health post) or “mobile” (>15km away from the health post). Outreach strategies vary based on the local community and available infrastructure. Microplanning led by head nurses, CHWs, and community actors helps determine whether fixed, mobile, or advanced strategies should be implemented to reach most children and how often each category is required to ensure high coverage.

#### 3.5.3. Data used for decision-making through coordination meetings

Coordination meetings increased transparency of immunization program performance across subnational levels and created an incentive for improvement via peer-to-peer engagement. Nearly all key informants at the national, regional, district, and health post levels described coordination meetings as vital to improvements in data monitoring and evaluation.

The National Health and Social Development Plan 2019-2028 increased the frequency of coordination meetings and supportive supervision to improve accountability, transparency, and support in intervention implementation, as illustrated in Table 4 [36]. With the increased frequency of coordination meetings, the government and partners at the national level can work directly with the regional and district levels to review data and performance using feedback bulletins and the DHIS2. The national-level annual coordination meeting is chaired by the Minister of Health and brings together regional and district level physicians and external partners to review immunization performance indicators, follow up on recommendations from the previous year, discuss lessons learned, identify solutions to frontline challenges, and determine next steps. At the health post level, stakeholders also monitor activities and conduct community feedback sessions where they share results from monthly coordination meetings.

**Table 4.**
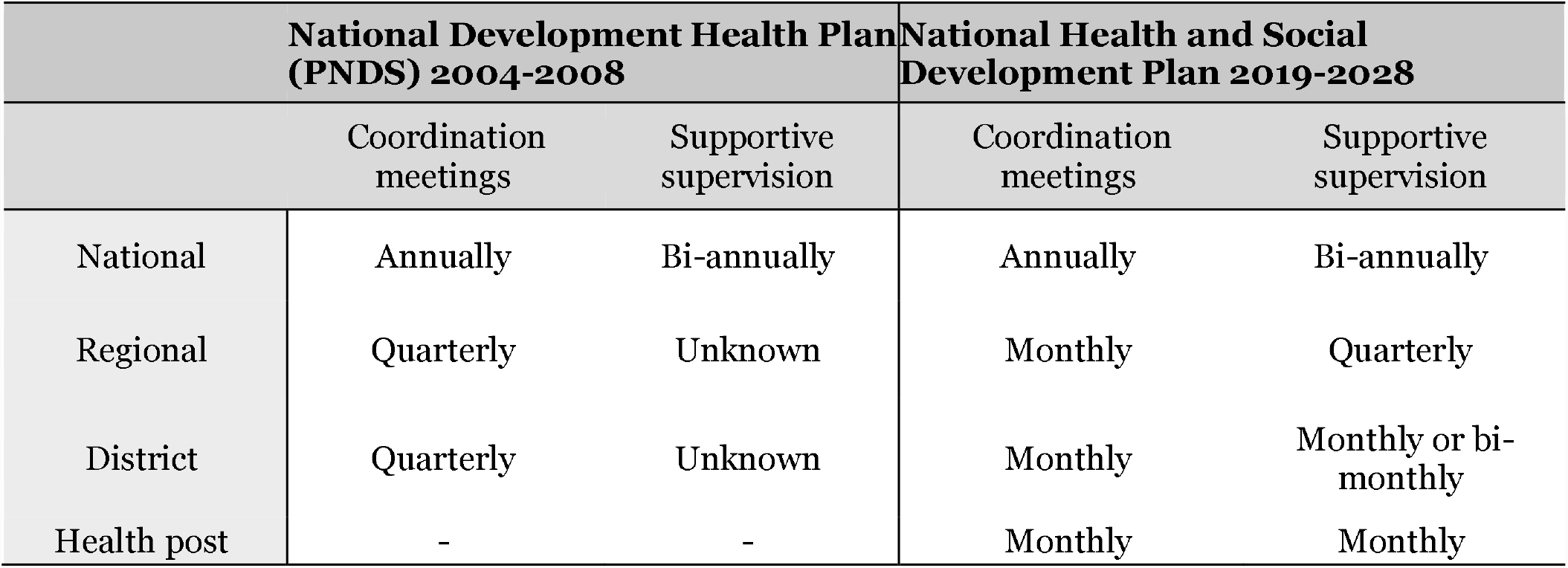
Increase in the frequency of coordination meetings and supportive supervision.

> *“I remember when there was the annual review of the EPI, and it was chaired by the cabinet*… *to discuss vaccine performance. That left an impression on me because I was the chief doctor of a district, and it was in front of all our peers that we had to justify poor performance while the best was congratulated*… *The minister himself came to preside until 2018, and that really sparked this motivation of the actors. There is also the publication of the feedback bulletin that we receive every month*… *These are things, in my opinion, that really marked a turning point in the management of the program.” (External partner at the national level)*

### 3.6. Community ownership mechanism

#### 3.6.1 Community health development committees support local health programs

Community health development committees (*Comités de Développement Sanitaire*, referred to as CDS) play a significant role in the implementation, management, financing, and decision-making for health programming. The CDS functions at the community level and includes neighborhood delegates, mayors, religious leaders, teachers, and other community stakeholders. Each CDS works with their corresponding health post and fosters collaboration between local decision-makers and the health sector (through nurses and community health workers) to support community health improvements and initiatives. The CDS collects health post revenue from the health post, holds and manages the community funds, and provides resources for the implementation of activities, which may include purchase of vehicles or fuel for transport, cold chain maintenance, improvements to the health post infrastructure, consumables, training resources, or materials for campaigns and outreach. Figure 6 illustrates the planning and management of funds in Senegal.

**Figure 6.**
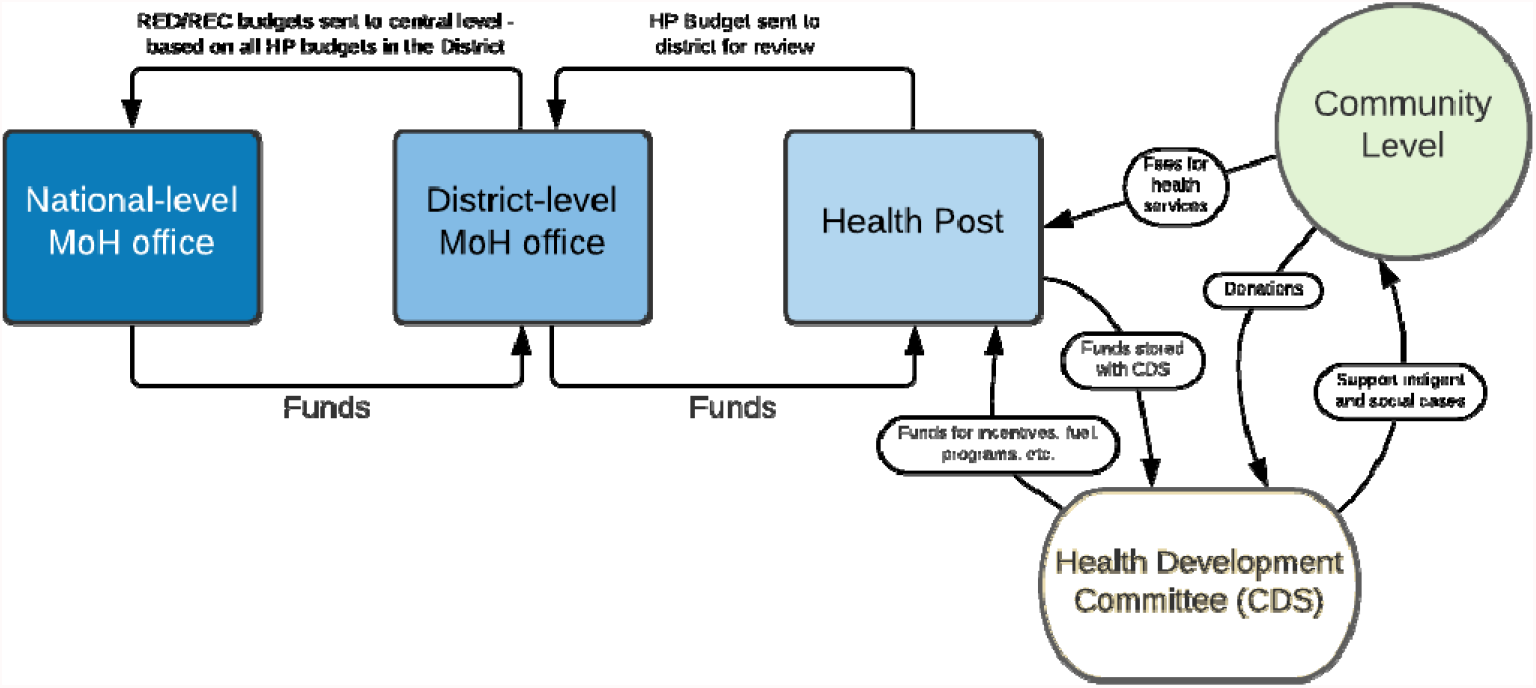
In-country planning and management of funds for immunization in Senegal.

The role of the CDS was elevated by the national-level government and is now considered the lowest administrative unit within the government system. The purpose of this decision was to promote community ownership of health and development outcomes as the national-level government determined that individuals within the community are most suited to identify their own needs and develop strategies for addressing them [37]. Each CDS facilitates the adaptation of vaccine programming to meet local and context-specific needs by providing a platform for meetings and supporting community-level planning. To prioritize equitable health outcomes, about 5% of CDS revenue is allocated to “support indigent and social cases” as of 2018 [36]. During district-level coordination meetings, CDS representatives are asked to share success and challenges within their communities which support peer-to-peer learning.

#### 3.6.2. Community health workers bridge communities to the health system

Volunteer community health workers (CHWs) spearhead the promotion and demand generation of vaccines and play a crucial role in supporting service delivery. CHWs support head nurses (at the health post level - who administer the vaccines), organize outreach services, remind caregivers of vaccine activities and vaccines needed, and find dropouts. CHW programming supports community ownership by assigning the responsibility of health-related activities and outcomes to volunteers working with the communities they serve.

Five cadres of CHWs contribute to vaccine delivery and uptake through curative, preventative, and promotive care. However, *relais, bajenu gox* (“godmothers”), and home health workers (DSDOMs) are the three cadres that are primarily involved with vaccine programming. These cadres conduct educational activities with caregivers, discuss the benefits of vaccines and dispel misinformation, collaborate closely with community members to plan and coordinate vaccine events, and provide personal support to caregivers. The health system created *bajenu gox* and DSDOM positions in 2013 and 2008, respectively, to address a gap in human resources. Community members, including parents, described their trust in CHWs, which supports effective health messaging and demand generation. Bajenu gox raise awareness among family members of different generations and genders; as elders, their words and advice are respected. Other CHW cadres promote routine immunizations through ante-natal care and coordinate community health events.

> *“Community health workers play a very important role in what we call the Senegalese community health system. Community relais are in direct contact with the population. They are the ones who live in the community, and they identify more with the community. So, there is a relationship of trust between community relais and the population, hence their auxiliary role. I would say they are health auxiliaries who help a lot in raising awareness, but also help in the acceptability of health policies by the population. I think that community relais play a fundamental role, and today, I think that COVID-19 has demonstrated it. At a certain point, when we felt reluctance from the population, we turned to*…*the bajenu gox, the community relais, the [health] journalists who all really played a fundamental role in what we call community engagement.” (National level government stakeholder)*

CHWs were motivated by tangible and intrinsic incentives, including money, food, assistance in tending crops, recognition for their commitment and accomplishments, public accolades, and certificates. CHWs also valued the training and capacity building, which increased their self-efficacy. Selection by the community translated into a strong sense of responsibility to their position and the health of children. However, CHWs and health facility staff expressed frustration at the government’s lack of budget provision for CHWs, leaving the CDS to determine and pay for incentives.

#### 3.6.3. Community engagement generates demand for vaccines

All key informants, from the national to community level, reported that community demand for vaccines and knowledge about the benefits of vaccines and how to access them was critical for improving routine immunization coverage. In addition to the work of CHWs, demand generation and public awareness were accomplished through policy development from the MoH, media engagement, involvement of community actors, and school outreach activities. Community actors involved with demand generation activities are described in Table 5.

**Table 5.**
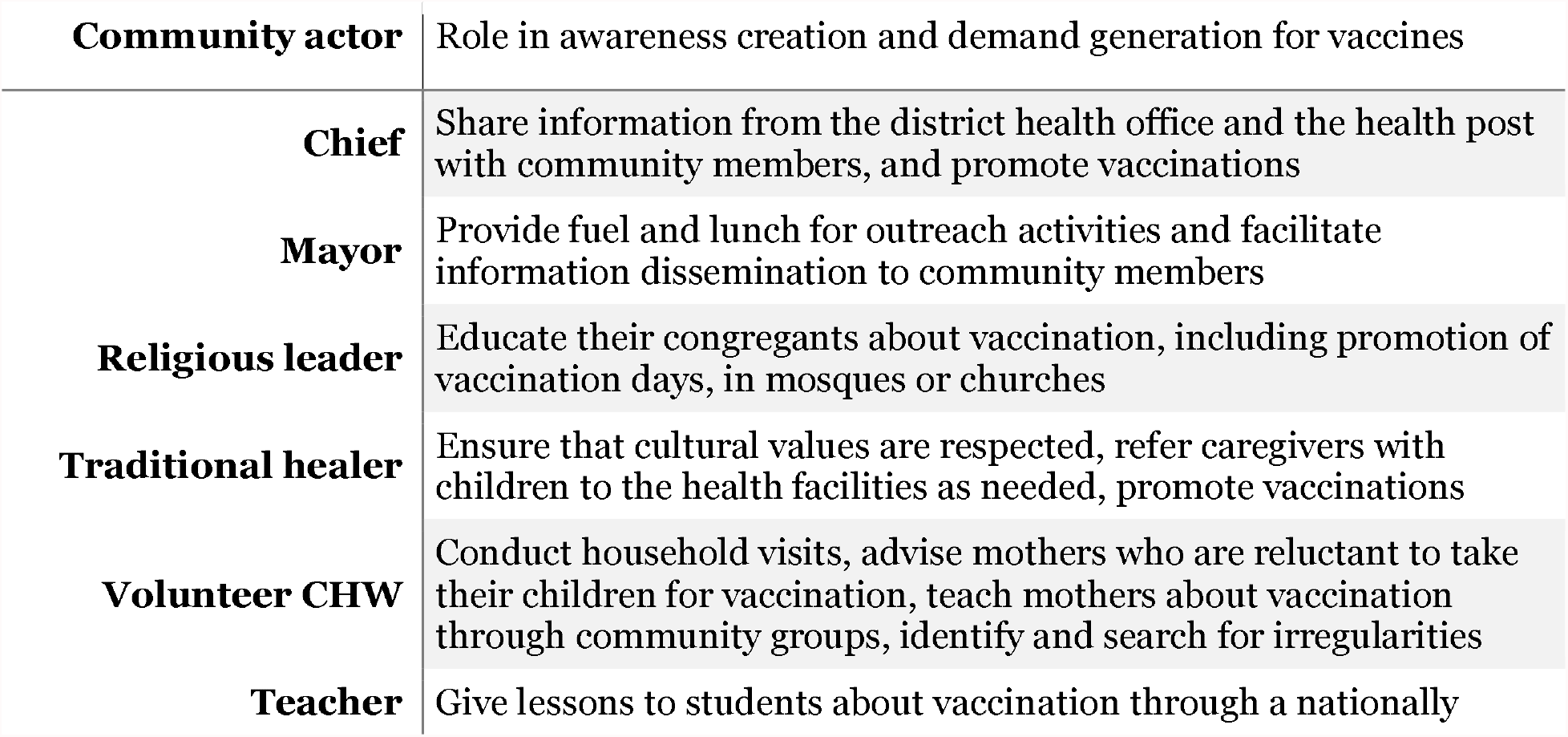
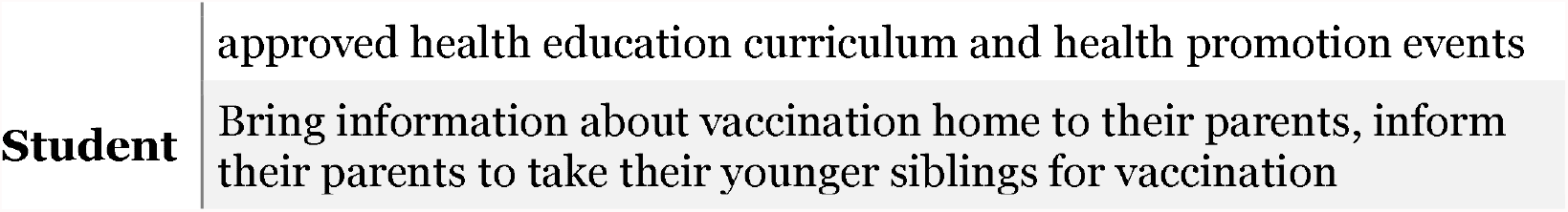
Community actors and their role in generating demand for vaccines.

In Senegal, community health programming began with the Bamako Initiative of Public Health which was fully adopted in 1991 [38-40]. The Bamako Initiative identified community participation as fundamental for the success of primary health care. Key informants spoke to the importance of national-level support for community engagement policies and programming in order to effectively coordinate with community leaders, who may act as gatekeepers to their communities. Regional development committees within the MoH rely on coordination with community actors and representatives from community-based organizations. Community actors are invited to planning sessions to support the organization of immunization campaigns and travel door-to-door on vaccination days in order to mobilize mothers to bring their children for vaccination. Community engagement is tailored depending on the community; for example, one key informant mentioned the mobilization of local youths to spread information in their communities and remind mothers about vaccine appointments on the day.

District health officers and health post staff provide chiefs and religious leaders with information about vaccination, which they share with community members among hard-to-reach populations. Religious leaders generate public awareness through education to their congregations, either before or after religious activities or events, in community mosques or churches. Religious and traditional leaders ensure that cultural values are respected, and therefore gain more respect and authority in their communities [41].

### 3.7. Equitable access to vaccine services and promotion

While our objectives were to focus on why Senegal is an exemplar in vaccine delivery, our data also suggests that diverse populations within the country may receive inequitable services. While national-level coverage in Senegal is high, regional coverage has greatly varied; it is critical to address gaps in the vaccine delivery system to meet the Constitutional promise of “health as a human right”. Ongoing efforts to address equity concerns include: 1) Overcoming geographical barriers through health post expansion and outreach services supported by national-level policy and external partner funding; and 2) Addressing social, economic, and cultural factors through tailored messaging and local ownership of vaccination programming. Challenges and solutions related to achieving equitable access to vaccines in Senegal, as reported by key informants, are illustrated in Table 6. For example, implementing outreach services to address lack of access in remote areas, scheduling vaccination services to align with community availability, and tailoring strategies for disseminating health information to circumvent challenges with female literacy. Solutions included in the table below have not been implemented country-wide, should be tailored locally/regionally, and show potential to reduce social and environmental inequities.

**Table 6.** Equity of vaccine services and promotion – Challenges and Solutions in Senegal.

| Challenge | Solution(s) |
| --- | --- |
| Difficult to access health services and facilities in rural areas | Health post expansion |
|  | Targeted outreach services |
| Difficult for health workers to access vaccination and health care recipients | Budget for appropriate transportation and gas |
|  | Investment in improved infrastructure |
| Caregivers unable to access health facilities for vaccines at set times | Adjust time or day when vaccination services are offered |
| Lack of resources for assisted delivery and ante-natal care | Health post and health hut expansion |
|  | Ante-natal care and education |
| Female illiteracy acts as a barrier for spreading public awareness so mothers so that they can make informed decisions about health | Oral education and messaging |
|  | Visual messaging |
|  | Increased enrollment and retention of female students |
|  | Verbal reminder system for vaccination days, vaccinations needed (face-to-face, phone calls) |
|  | Increased outreach to fathers for support and engagement for mothers and child health |
| Gender inequity may reduce mother's power in decision-making related to health | Targeted outreach to fathers on health decision-making partnerships |
| Nomadic populations' movement hinders access to immunization days, outreach, and impedes efforts for tracking/monitoring | Increased engagement with extended community for education and outreach, through existing leadership |
|  | Communication between head nurses for tracking |
| Vaccine hesitancy among minority/marginalized populations | Engage CHWs that speak specific languages |
|  | Engage CHWs from minority populations |
|  | Work with community leaders to promote acceptance |

### 3.8. Implications for policy development, strategic planning, and investments in vaccine programming for other LMICs

Findings from this research may have implications on vaccine policy, programming, and investments in other low- and middle-income countries. Though some of the strategies described in this paper are unique to the Senegalese context, many of the highlighted success factors may have salience in other settings or apply to other health systems for a horizontal approach to healthcare. Namely, collaboration and effective communication at multiple levels of government were crucial for the success of the vaccine delivery system in Senegal, a finding that applies beyond vaccine delivery to the entire health system. Our data highlights the importance of a more systemic assessment of health delivery systems, including important governance structures and processes for effective collaboration between stakeholders, both internal and external to the government.

We found that many improvements in immunization coverage emerged from adjacent successes in health systems strengthening (HSS). HSS has become a priority in recent decades as disruptions due to natural disasters, disease outbreaks, or civil unrest have exposed weaknesses in health systems [42, 43]. The Immunization Agenda 2030 includes a strong focus on HSS, building on GVAP, and states that sustainable immunization programming should be embedded within primary health care [1, 44, 45]. In Senegal, the government executed parallel improvements in HSS and vaccine delivery through health post expansion, training of health personnel and volunteers at all levels, improvements in data management and surveillance, and overall capacity building for health posts, health huts, and infrastructure required for outreach services. HSS was supported by the application of international guidelines and adaptations to address context-specific challenges and priorities.

Our data also suggests that a deliberate and consistent focus on community engagement and ownership of health programming is essential to the success of a vaccine delivery system, which aligns with existing literature [1, 46, 47]. In Senegal, community ownership was fostered through national policies and resources for capacity building, management of funds at the health post level by community health development committees, engagement of local leadership including mayors, chiefs, and religious leaders, and strength of the CHW program. Adapting service delivery through community involvement to address the needs of end-users also improved access to vaccines and made the system more resilient to challenges, including the COVID-19 pandemic.

The experience in Senegal suggests that the success and sustainability of the immunization program are supported by strong governance, collaboration, evidence-based decision-making, community ownership, and an overall commitment to health and prioritization of vaccine programming from all stakeholders and government officials. Overall, the strategies we identified align with the WHO Comprehensive Framework of Strategies and Practices for Routine Immunization, which focus on managing the immunization program (e.g., political commitment, partnerships), mobilizing communities, and generating demand, and monitoring progress [1]. This research more comprehensively examines *how* these strategies are operationalized to improve coverage. Findings from this research point to the need for programmers and policymakers to better understand the strengths and limitations of the underlying governance structures that these strategies rely on.

## 4. Limitations

This study has several limitations. First, we focused on Senegal as a positive deviant in vaccine delivery but were unable to carry out a similar analysis in a non-exemplary country to compare immunization coverage and vaccine delivery systems. Second, the research tools focused on the factors that drove catalytic change and did not focus on interventions or policies that were unsuccessful. Third, using qualitative methods to understand historical events was challenging; interviewees often spoke about current experiences rather than discussing historical factors. However, research assistants probed respondents to reflect on longitudinal changes in the immunization program. Fourth, some policy documents – including national-level strategic plans - were not available for our review. And finally, the COVID-19 pandemic had significant impacts on data collection as national-level stakeholders were unavailable for interviews due to the prioritization of the pandemic response and community-level FGDs were limited due to safety precautions.

## 5. Conclusion

Our research revealed that success factors related to collaboration and community involvement led to improved immunization coverage in Senegal. We conducted a theory-informed investigation into the historical drivers of *how* and *why* Senegal achieved catalytic growth in early childhood vaccine coverage. The vaccination program in Senegal was supported by evidence-based strategic planning at the national level, alignment of priorities between governmental entities and external partners, and strong community engagement initiatives that fostered local ownership of vaccine delivery and uptake.

High and sustained routine immunization coverage was likely driven by prioritization of the EPI, improved surveillance systems, a mature and reliable community health worker program, and tailored strategies for addressing geographical, social, and cultural barriers. These findings – along with the approach used to understand underlying strengths of the vaccine delivery system – could be adapted to other contexts and contribute to improvements in national vaccine delivery systems.

## Data Availability

The data are not publicly available as all data are confidential. Data may be available upon request.

## Acknowledgements

We thank the Institut de Recherche en Santé de Surveillance Epidemiologique et de Formation (IRESSEF) in Dakar, Senegal for their partnership in this study. We gratefully acknowledge the participants who gave their time and insights to help us better understand Senegal’s vaccine delivery system, along with facilitators from the Ministry of Health and Social Action for supporting this research, especially Dr. Ousseynou Badiane and Dr. Abdoulaye Mangane. In addition, we thank Sarah Chesemore, Anna Rapp, Tove Ryan, and Ethan Wong from the Bill and Melinda Gates Foundation; Kate Buellesbach, Nancy Fullman, Nathaniel Gerthe, Gloria Ikilezi, Caitlyn Mason, David Phillips, and Oliver Rothschild, Jordan-Tate Thomas, and Angela Wang from Gates Ventures; and the Vaccine Exemplars Research Advisory Group for their insights, specifically Agnes Binagwaho, Laura Craw, Carolina Danovaro, Anuradha Gupta, Heidi Larson, Penelope Masumbu, Kate O’Brien, Helen Rees, Lora Shimp, and Aaron Wallace.

